# Single-dose epidural bupivacaine vs placebo after lumbar decompression surgery: A Randomized controlled trial

**DOI:** 10.1101/2023.08.20.23294347

**Authors:** Sem M. M. Hermans, Aniek A. G. Lantinga-Zee, Ruud Droeghaag, Henk van Santbrink, Wouter L. W. van Hemert, Mattheus K. Reinders, Daisy M. N. Hoofwijk, Sander M. J. van Kuijk, Kim Rijkers, Inez Curfs

## Abstract

**Introduction:** Adequate postoperative pain management following lumbar spinal decompression surgery is important as it will lead to early mobilization, less complications and shorter hospital stay. Opioid consumption should be limited due to their frequently accompanied side effects and their addictive nature. During the final phase of lumbar decompression surgery, the epidural space becomes easily accessible. This might be an ideal moment for surgeons to administer an epidural bolus of analgesia, as a safe and effective method for post-operative pain relief.

**Methods:** This is a double blind randomized controlled trial comparing a single intraoperative bolus of epidural analgesia using bupivacaine 0.25% to placebo (NaCl 0,9%) and its effect on postoperative pain following lumbar spinal decompression surgery. The primary outcome was the difference in NRS pain between the intervention and placebo groups during the first 48h after surgery. It was hypothesized that the intervention group will have lower postoperative NRS pain scores.

**Results:** Both the intervention group and the placebo group consisted of 20 randomized patients (N=40). We observed statistically significant lower NRS pain scores in the intervention group in comparison with the control group, with a difference of −1.9 (±1.1). The average pain score was lower in the intervention group at all postoperative time-points. Opioid consumption, quality of life and satisfaction were similar between study groups. No study related complications occurred, and complications rate did not differ between study groups.

**Conclusion:** This randomized controlled trial shows that administrating a bolus of intraoperative epidural bupivacaine is a safe and effective method in reducing early postoperative pain following lumbar decompression surgery.

## INTRODUCTION

Neurogenic claudication due to lumbar spinal stenosis is one of the most common degenerative spinal disorders.^1^ The stenosis is typically a result of spondylosis of the lumbar spine which narrows the spinal canal. Lumbar spinal stenosis mostly affects individuals over the age of 60 years.^2^ The incidence of degenerative spinal disorders is rising along with the ageing of the population, leading to an increase in the number of lumbar decompression surgeries.^3^ Neurogenic claudication due to lumbar spinal canal stenosis is usually surgically treated. The most common procedures are laminectomy and interlaminar decompression. The goal of these procedures is symptom relief, alleviating complaints of pain, numbness and weakness of legs and buttocks.^4^

Adequate postoperative pain management is important as it will lead to early mobilization, less complications and shorter duration of hospital stay.^5^ Opioids are often used to attain these goals. Opioid consumption is frequently accompanied by unwanted side effects, such as respiratory depression, constipation, and nausea, which might influence mobilization and length of hospital stay.^6^ Moreover, patients tend to continue the use of prescribed postoperative analgesics, including opioids, after discharge because of their addictive nature.^7,8^ This will worsen the current societal problems related to chronic opioid use in Western countries.^9,10^

During the final phase of lumbar decompression surgery, the epidural space is exposed and easily accessible for administration of an epidural bolus of analgesia. This can be a safe and effective method for immediate post-operative pain relief. Current literature on epidural administration of intraoperative analgesics in lumbar decompression surgery is heterogeneous. No consensus exists concerning the optimal technique for postoperative pain reduction. Several administrative methods and agents can be considered, including steroids.^11–15^ Although epidural administration of steroids has the potential to reduce postoperative pain in patients undergoing lumbar decompression surgery, there are several concerns regarding its use.^13,14,16^ The main concern is the increased risk of surgical site infection and possibly epidural abscesses.^17,18^ According to some studies, non-steroidal analgesics (i.e. amide, morphine) administrated to the epidural space may similarly achieve postoperative pain reduction, without the increased risk of infections.^19–23^

Controversy remains regarding the effectiveness and safety of intraoperative epidural application of analgesics. Therefore, an RCT comparing epidural analgesia using bupivacaine 0.25% to placebo (NaCl 0,9%) was designed to investigate the added value of an intraoperative epidural non-opioid analgesic. Results of this study will provide evidence-based treatment recommendations.

## METHODS

### Study design

This was a prospective, double blind, randomized controlled trial (RCT). Patients, clinicians, researchers and the statistician were blinded. Only the pharmacy personnel were not. Recruitment took place between 4 December 2020 and 16 January 2022. This study was registered in the Dutch National TrialRegistry (registration number: NL8030; currently transferred to the International Clinical Trial Registry Platform, ICTRP, https://trialsearch.who.int/Trial2.aspx?TrialID=NL8030) and was written in accordance with the Consolidated Standards of Reporting Trials (CONSORT) 2010 statement.^24^ Ethical approval has been granted by the Medical Ethical Committee Zuyderland, Heerlen, the Netherlands (registration number: Z20190138).

### Study population

Adult patients referred to the neurosurgical or orthopedic outpatient clinic who were candidates for open lumbar spine decompression surgery (laminectomy and interlaminar decompression) were eligible to participate in this RCT. Neurosurgeons and orthopedic surgeons referred these eligible patients to the researchers (SH and AL). The researchers informed the patients. Patients were included after informed consent in writing was obtained.

Patients with any of the following criteria were excluded from participation in this study: preoperative opioid use, previous radiotherapy at the intended surgical level, (progressive) motor failure and/or anal sphincter disorders which urges instant intervention, active spinal inflammation/infection, immature bone (ongoing growth), pregnancy, contra-indications for the use of bupivacaine or other amide-type local anesthetics, and/or inadequate command of the Dutch language.

### Interventions

After receiving antibiotic prophylaxis, the patient was brought under general anesthesia. All patients were intubated and then positioned in prone position. Short-acting opioids like sufentanil or fentanyl were used during surgery. To prevent the influence of long-acting opioids (e.g. morphine and piritramide) on the postoperative pain and therefore obscuring the result of the epidural analgesia, no morphine was administered during or at the end of surgery. A midline approach was performed, exposing the posterior lumbar elements including the facet joints. The spinal canal was decompressed through a laminectomy or interlaminar decompression. At the end of surgery, an epidural catheter was directly placed under the most superiorly exposed lamina and placed at minimum 5 to maximum 10 cm upward, depending on the level. The spreader was removed, and the wound was thoroughly irrigated with saline and closed. After closure of the fascia, 5 cc of the study solution, which was prepared and blinded by the pharmacy, was applied to the catheter. Either bupivacaine 0.25% (intervention) or NaCl 0.9% (placebo) was administrated. The catheter was removed immediately after administration of the bolus. The wound was closed without suction drainage. Patients who signed informed consent but were not eligible for randomization (dural tear or inability to place catheter) received the standard of care. Postoperatively, patients were transported to the recovery room, where they were monitored for a least one hour.

### Outcomes

The primary outcome was the average difference in NRS pain (0 - 10, 10 being ‘worst pain imaginable’) between intervention and placebo during the first 48h after surgery. NRS pain measurement was specific to the surgery site. With interval measurements at recovery entry, recovery exit, 2, 4, 6, 24 and 48 hours postoperatively. Cumulative opioid use was also monitored at recovery, 2, 4, 6, 24 and 48h postoperatively. Patients were able to receive additional analgesia until NRS ≤ 3. Health related quality of life and patient satisfaction were measured using EuroQol-5D (EQ-5D) and General Surgery Recovery Index (GSRI) to determine whether epidural analgesia leads to a better outcome than placebo. Patients filled out the questionnaires 24 hours after surgery. Adverse events such as dural tear, postoperative infection, deep venous thrombosis, hematoma, neurological deficits and other medical complications such as pneumonia, urine retention or urinary tract infection were also monitored up to 30 days following surgery. Length of hospital stay (days) was evaluated as time between hospital admission and discharge.

### Randomization and blinding

Allocation took place at the end of surgery after the epidural catheter was inserted. In case of a dural tear or inability to place the epidural catheter, patients were referred to the cohort group who received standard care. The pharmacy prepared blinded syringes with either bupivacaine or NaCl. The pharmacy marked the syringes with a kit-number (1,2,3 etc). These numbers corresponded with a computer-generated randomization list which was stored by the pharmacy. Syringes were prepared once a week under aseptic conditions, and stored at the pharmacy. They were collected at the end of the day, one day before a study patient was operated on. Upon leaving the pharmacy, syringes were stored at the OR complex at room temperature for maximally 24 hours. When a patient was eligible for randomization (placement of catheter had been successful) the first successive syringe was used. The surgeon noted the kit-number in the patient’s electronic file. Once the study was finished, the randomization list was unblinded by the pharmacy to clinicians, researchers, statisticians and patients.

### Statistical methods

The primary endpoint is the difference in pain between the intervention group and the control group. The sample size calculation was based on the difference between the intervention and control group in NRS at recovery entry and exit following surgery. Own historical data showed a standard deviation of 2 in NRS scores. A two-point reduction on the eleven-points (0 to 10) NRS pain score was considered clinically relevant.^25,26^ In order to obtain a clinically meaningful effect with 80% power, we needed 16 patients per group. When taking into account a 20% loss to follow-up, 40 patients (20 patients per group) were enrolled in this study.

Frequency tables were provided for all categorical demographic information. Continuous variables were presented as mean ± standard deviation (SD) or median ± interquartile range (IQR) depending on the distribution of the data. Analysis was performed by principal investigators using IBM SPSS statistical software package version 27 (SPSS Inc., Chicago, IL). Missing values were imputed using stochastic regression imputation using full conditional specification. Data was tested for normal distribution. When data was normally distributed the independent-samples t-test was used to determine statistical difference between groups (e.g. NRS pain score, cumulative opioid use). In case of absence for normal distribution, Wilcoxon’s Signed-Rank test was used. In addition, the group differences over time were determined (e.g., slopes of the relation between time and pain) using a linear mixed-effects model with a random intercept on patient-ID and slope of time. The model included group and time as fixed factors, and the interaction between group and time. Categorical data was compared between groups using Pearson’s Chi-Square test and Fisher’s exact test. A p-value ≤ 0.05 was considered statistically significant.

## RESULTS

### Study population and surgical characteristics

Patients’ characteristics are described in Table 1. A total of 44 patients were included in this RCT. Four patients were not randomized due to a dural tear intraoperatively and thus received standard care. Twenty patients were randomized to the intervention group (bupivacaine), and 20 patients to the control group (placebo NaCl). All of these patients received the allocated treatment as per protocol. Surgical characteristics are described in Table 2. There was a significant difference in blood-loss between the intervention and control group (p=0.008). Two complications occurred in the first 30 days following surgery. There was no statistically significant difference in rate of complications between the study groups.

### Primary outcome

Baseline NRS pain score did not statistically significantly differ between study groups (p = 0.115). Statistically significant lower mean NRS pain scores were observed in the intervention group in comparison with the control group postoperatively (−1.9 (±1.1), p=0.013). No statistically significant effect was observed for time (p=0.119) and group-differences also did not significantly change over time (time*group-interaction, p=0.113). In Figure 1 the pre- and postoperative NRS pain score is outlined for both study groups.

**Figure 1:**
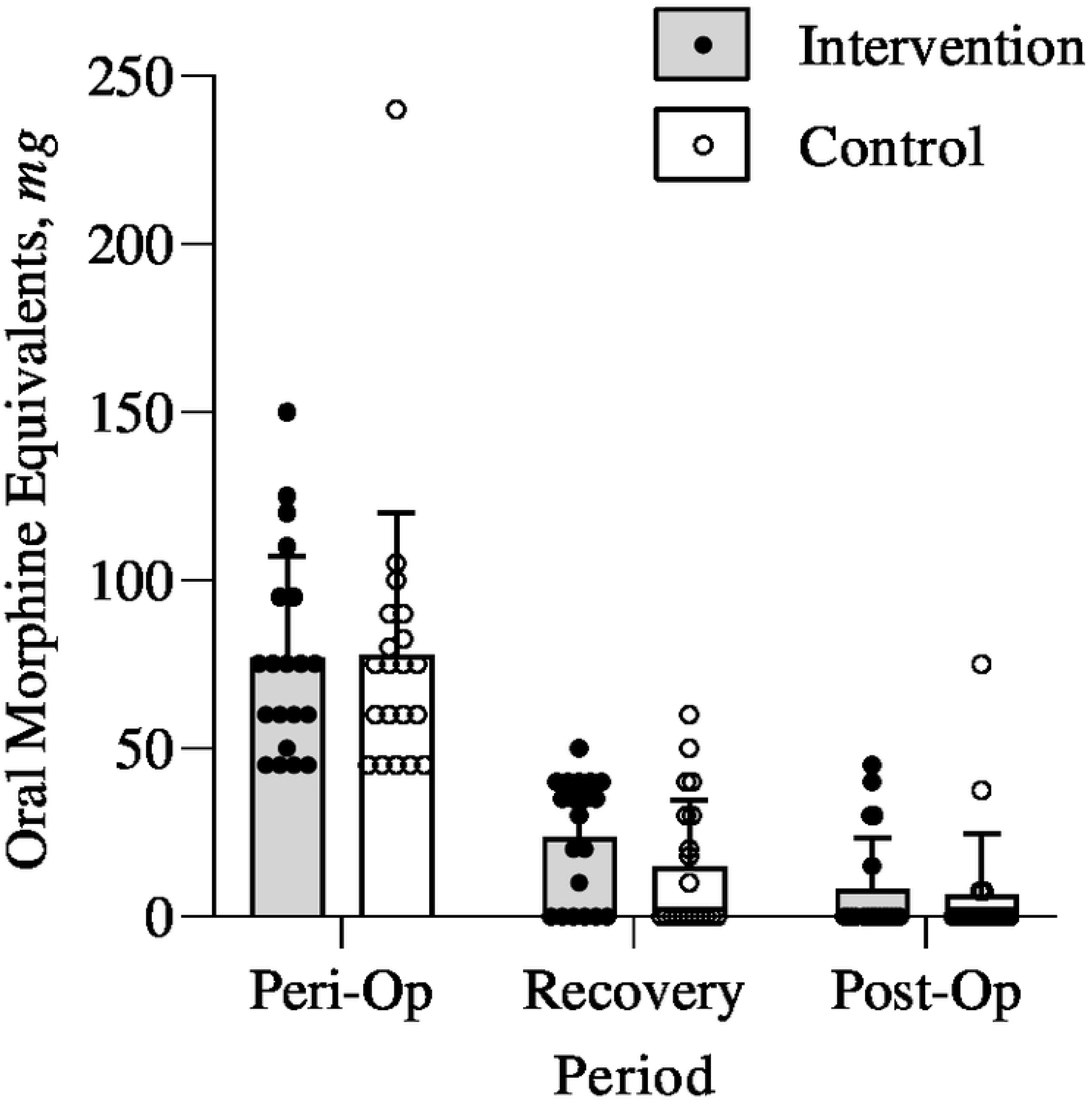
Pain score pre- and postoperative. NRS pain score is presented as means with SD. Abbreviation: OP= operative.

### Secondary outcome

Perioperative opioid use was not statistically significantly different between intervention and control group (Figure 2). Thirty-eight patients (95%) received sufentanil, one patient received alfentanil (3%), one patient received remifentanil (3%), and six patients also received piritramide (15%). Postoperatively, 28 patients received piritramide (70%), five patients received oxycodone (13%), and two patients received morphine IV (5%).

**Figure 2:**
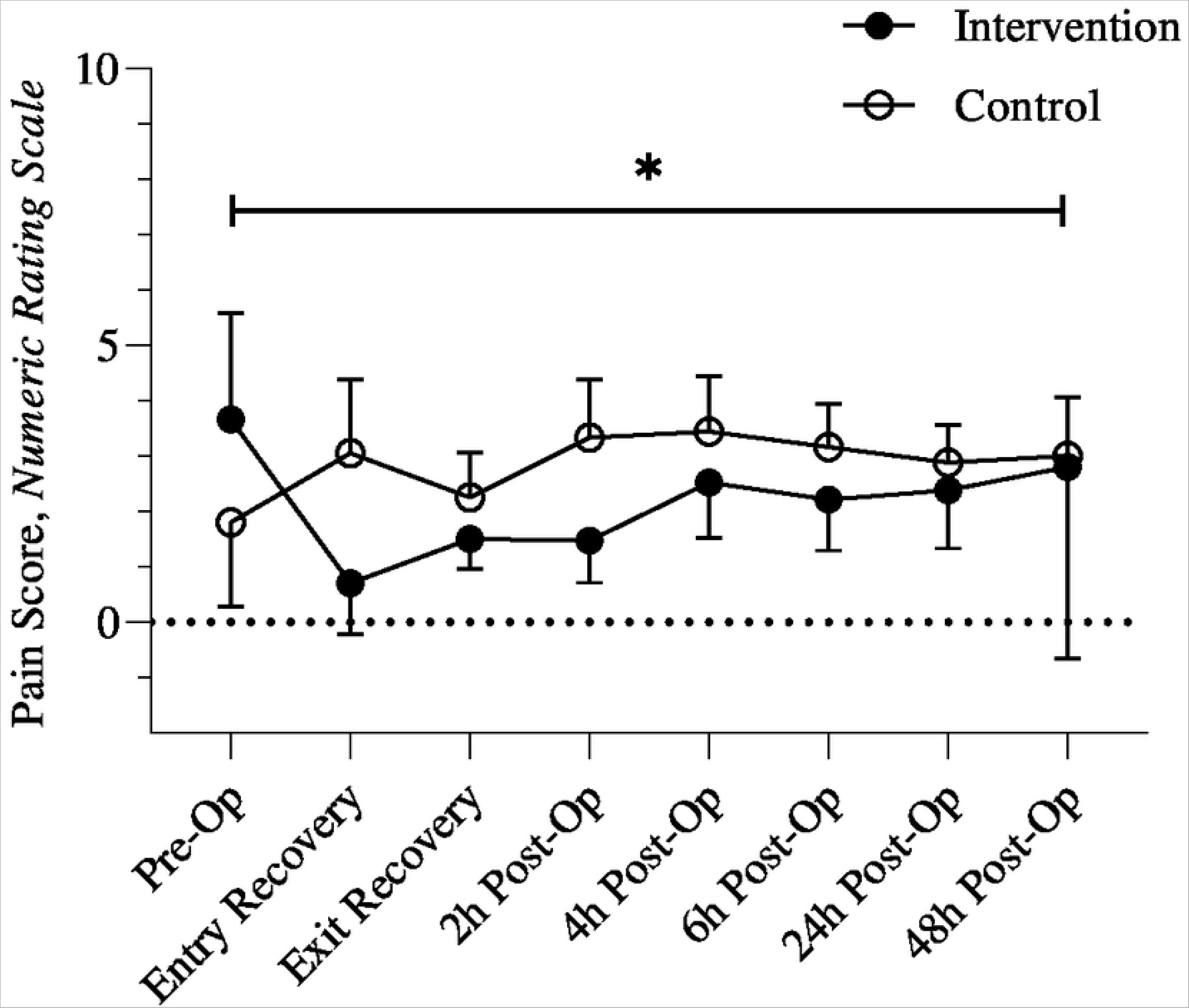
Peri- and postoperative opioid use. Morphine equivalent is presented as means with SD in mg. Abbreviation: OP= operative.

EQ-5D and GSRI outcomes at 24 hours postoperatively did not reveal any differences between study groups. EQ-5D score was 0.655 and 0.717 and GSRI was 70.18 and 63.50 for intervention group and placebo group, respectively.

### Standard care

Four patients received standard care, as randomization for either the intervention or control group did not take place due to a perioperative dural tear. In this group, two patients (50%) were diabetic. None of these four patients had a history of spine surgery, and none smoked. There were no relevant differences in baseline or surgical characteristics between the four patients that received standard care and the other two groups. Besides the four dural tears, no complications occurred in the standard care group.

Mean preoperative NRS was 0 (N=2). The mean NRS score at the time of entrance at recovery was 2.5 (N=4), and 2.3 (N=4) upon departure from the recovery. At 2, 4, 6, and 24 hours postoperatively, the mean NRS scores were 2.0 (N=2), 1.5 (N=2), 0.3 (N=3), and 0.3 (N=3), respectively. NRS at 48 hours postoperatively was not available for any of these patients.

Mean EQ-5D and GSRI 24 hours postoperatively was 0.834 (N=4) and 76.50 (N=4), respectively. The peri-operative opioid use was the equivalent of 191mg (±234mg) orally administered morphine. Opioid use at recovery was 28mg (±18), and the opioid use within the first 48 hours postoperatively after discharge from recovery was 0mg.

## DISCUSSION

This study gives new insight in the value of intraoperative epidural analgesia using bupivacaine compared to placebo following lumbar decompression surgery. The most important discovery is that immediate post-operative pain scores are reduced with bupivacaine. The average pain score during all postoperative hours was low to moderate (NRS < 3.5) in all study groups. Because of these low values, the difference in pain scores between study groups appears minimal. However, at entry of the recovery and 2 hours postoperatively NRS of the control group is twice as high as the intervention group. In the intervention group, a reduction of >2 NRS points is observed at the first three postoperative timepoints, which is not the case in the control group. The difference in pain scores between intervention and control group decreases at later postoperative hours. This may be explained by the fact that the effect of the bupivacaine bolus is worn off, as the elimination half-life is approximately three hours.^27^

No differences are observed in total opioid consumption between study groups in the postoperative period. Although we expected that total opioid consumption would be lower in the intervention group compared to the placebo group, it should be noted that equal opioid consumption across groups is established with lower reported pain in the intervention group. This shows that better pain control is achieved in the intervention group while consuming the same number of opioids. Additionally, the sample size of the current study is calculated based on difference in pain scores between intervention group and control group. Possibly, a different sample size is required to investigate the difference in postoperative opioid consumption after epidural bupivacaine administration. This might explain the discrepancy between our current findings and the systematic review and meta-analysis we previously performed regarding intraoperative epidural analgesia for pain relief after lumbar decompressive spine surgery.^28^ It is noteworthy that most studies investigating postoperative analgesia consumption describe a significant decrease in pain in the treatment group. However, only one study investigated postoperative opioid consumption, which dated from 1992.^29^ Other studies investigated tramadol or diclofenac consumption following surgery. Most of the included studies in our review implemented morphine as their analgesic agent. It is recognized that epidural administration of morphine increases the risk for urinary retention.^30^ Non-opioid analgesics, such as bupivacaine, might be the key to bypass complications linked to epidural analgesics administration. In the current study, not a single patient displayed symptoms of urinary retention. The complications observed in this study are related to wound healing, for which one patient received additional surgical intervention and antibiotic treatment. Wound healing related complications are inherent to surgical procedures and the numbers we report are in line with current literature.^31^ Several methods of delivering local or regional analgesia during spine surgery are described in current literature.^28^ In this study we choose to use a catheter to deliver the bolus as it has the possibility to be delivered more rostrally, resulting in a potential more immediate effect compared to sponges or direct application. Furthermore, a bolus rather than a continuous infusion was used as patients were encouraged to mobilize as soon as possible following surgery. Also, maintaining catheters in situ postoperatively increases the risk for infection.^32^

In the 24 hours following surgery, both EQ-5D and GSRI scores are high in all patients. These scores indicate that the majority of patients recovers well one day postoperatively and may also indicate that the difference in pain score across groups does not influence the perceived health-related quality of life and patient satisfaction. One can therefore wonder to what extent the different pain scores are clinically relevant. Nonetheless, this study was not powered on EQ-5D or GSRI, thus making assumptions based on these outcome data must be done with caution. The overall length of hospital stay was low, with a mean of 1.7 days across all study groups. This indicates that most patients left the hospital the second postoperative day. Current literature describes a mean length of stay of 2.1 days for patients following elective laminectomy.^33^ This small difference might be explained by intercontinental differences in studies.

Finally, in a small group of patients (N=4) a perioperative dural tear occured. These patients received standard care, as administrating an epidural bolus is deemed unsafe. In this group, the perioperative opioid use is higher compared to intervention and control groups. This is expected as exclusion from receiving a bolus of placebo or bupivacaine meant the patient was able to receive long-acting opioids during surgery. Since the standard care group consisted of four patients, statistical tests to compare this group with the other two groups is not deemed meaningful.

### Limitations

This study is bound by some important limitations. First, there is some missing outcome data. This is mainly the result of delayed or missed measurement moments. At the 48-hour measurement interval, the missing data is for the most part caused by the discharge of patients before measurement. One can suppose that NRS pain score was low, considering a patient is able to be discharged from the hospital. Percentage completed interval measurements were; recovery entry (100%), recovery exit (100%), two hours (84%), four hours (84%), six hour (91%), 24 hours (86%), and 48 hours postoperatively (27%). There were no statistically significant differences in rates of missing data between intervention and control group.

Second, a possible bias might have been introduced as some recovery nurses consistently administer opioids when patients return from OR, as they expect the patient to have pain. Our protocol describes that patients should only receive additional analgesia when NRS > 3. Not all personnel followed these guidelines at the start of the study.

Third, the primary outcome of this study is the NRS pain score, which is at risk for some sort of subjective discrepancies between patients. As is the case for all patient related outcome measurements used in this study, being the EQ-5D and GSRI. Postoperative opioid consumption is an objective outcome of this study, however no differences between study groups were observed. Further research to reduce opioid consumption is relevant, as opioid consumption should be limited after surgery, given its known side effects and growing societal problems related to chronic opioid use, especially in surgical patients.^9,10^ In the US the opioid epidemic was initially driven by increased consumption and availability of pharmaceutical opioids. However, an increasing number of opioid overdoses are now related to heroin and illicitly manufactured fentanyl and fentanyl analogs.^34^ Addressing this epidemic requires, among others, reducing opioid consumption in hospital setting. In some cases, it might be possible to omit opioids completely during an admission for lumbar decompression surgery. Ten patients (50%) in the treatment group and eight patients (40%) in the placebo group did not receive any morphine during their postoperative period. Patients should only receive opioids when pain is not adequately managed.

## CONCLUSION

This randomized controlled trial indicates that administrating a bolus of intraoperative epidural bupivacaine is a safe and effective method to reduce early postoperative pain following lumbar decompression surgery. This method of analgesia may be a useful adjunct in patients undergoing decompressive lumbar spine surgery.

## Data Availability

All relevant data are within the manuscript and its Supporting Information files.

https://trialsearch.who.int/Trial2.aspx?TrialID=NL8030

## TABLE LEGEND

**Table 1: Characteristics of included patients between intervention and control group.**

Abbreviation: ASA classification= American Society of Anaesthesiologists classification, BMI= Body Mass Index, SD= Standard Deviation

**Table 2: Surgical characteristics of included patients between intervention and control group.**

P-value <0.05 stands for statistically significant difference which is marked with *.

Abbreviation: SD= Standard Deviation

## ADDITIONAL FILES

Additional File 1 – Opioid conversion table

Additional File 2 – Table with NRS pain score difference adjusted for baseline pain.

